# Pain Reduction by Inducing Sensory-Motor Adaptation in Complex Regional Pain Syndrome (CRPS PRISMA): Protocol for a Double-blind Randomized Controlled Trial

**DOI:** 10.1101/19000653

**Authors:** Monika Halicka, Axel D Vittersø, Michael J Proulx, Janet H Bultitude

## Abstract

**Background:** Complex Regional Pain Syndrome (CRPS) presents as chronic, continuous pain and sensory, autonomic, and motor abnormalities affecting one or more extremities. People with CRPS can also show changes in their perception of and attention to the affected body part and sensory information in the affected side of space. Prism Adaptation (PA) is a behavioural intervention targeted at reducing attention deficits in post-stroke hemispatial neglect. PA also appears to reduce pain and other CRPS symptoms; however, these therapeutic effects have been demonstrated only in small unblinded studies. This paper describes the protocol for an ongoing double-blind, randomized, sham-controlled clinical trial that will evaluate the efficacy of PA treatment for CRPS. The secondary aims of the study are to examine the relationships between neuropsychological changes (such as spatial attention, space and body representation, and motor spatial performance) and clinical manifestations of CRPS, as well as symptom improvement.

**Methods:** Forty-two participants with upper-limb CRPS type I will undergo two weeks of twice-daily PA treatment or sham treatment. The primary outcome measures are current pain intensity and CRPS severity score, measured immediately before and after the treatment period. Secondary outcome measures include the results of self-report questionnaires about pain, movement, symptoms interference, and body representation; clinical assessments of sensory, motor, and autonomic functions; and computer-based psychophysical tests of neuropsychological functions. Data are collected in four research visits: four weeks and one day before treatment, and one day and four weeks after the end of treatment. Additional follow-up through postal questionnaires is conducted three and six months post-treatment.

**Discussion:** It is hypothesised that participants undergoing PA treatment, compared to those receiving sham treatment, will show greater reduction in pain and CRPS severity score, and improvements on other clinical and neuropsychological measures. Also, more pronounced neuropsychological symptoms are predicted to correlate with more severe clinical CRPS symptoms. This study will provide the first randomized double-blind evaluation of the therapeutic effects of PA that could be implemented as a rehabilitation method for CRPS, and will contribute to the understanding of how neuropsychological changes in body representation and attention pertain to the manifestation and treatment of CRPS.

## Background

People with Complex Regional Pain Syndrome (CRPS) experience continuous pain and a range of sensory, autonomic, and motor signs and symptoms. The condition primarily affects one or more extremities, which can become swollen and present with asymmetric changes in hair, nail and skin growth, sweating, limb temperature, and skin colour. Further clinical features of CRPS include allodynia (non-nociceptive stimulation perceived as painful) and hyperalgesia (mildly noxious stimulation experienced as extremely painful), as well as motor disturbance in the affected limb (e.g., decreased range of movement, weakness, tremor, and muscle contractions [2, 3]). Although CRPS usually develops after an injury to the limb (e.g., a fracture [4]), it can also develop spontaneously [5], and the symptoms are disproportionate to any inciting trauma [3]. There is no known cause of CRPS, however, several pathophysiological mechanisms are suggested to play a role in the development and maintenance of this syndrome, including neuroinflammation, nociceptive sensitization, vasomotor dysfunction, and maladaptive neuroplasticity [2].

CRPS patients have shown reduced attention to tactile [6–8] and visual stimulation on the affected limb and in external space near it [9, 10]. These biases appear to be associated with the side of space in which the limb usually resides [7, 9] rather than a tendency to pay less attention to the affected body parts per se. These space-based attention changes resemble those found in post-stroke hemispatial neglect patients [11].

One emerging treatment for CRPS is Prism Adaptation (PA). PA is a form of a sensory-motor training used to reduce lateralised attention deficits in post-stroke hemispatial neglect. The treatment involves performing a pointing task while wearing goggles fitted with prismatic lenses that induce a lateral deviation of the visual image. Due to this visual shift, patients’ pointing initially errs in the direction of prismatic displacement. With repeated movements, pointing becomes more accurate through an adjustment of pointing movements in the opposite direction to the optical shift, indicating a realignment of the sensory-motor reference frames [12, 13]. Once the goggles are removed, a negative after-effect is observed whereby pointing movements err in the opposite direction to the earlier optical shift. Using PA to induce pointing after-effects towards the neglected side reduces post-stroke hemispatial neglect [14–22].

The apparent attention bias in CRPS patients led to investigations of whether PA could also have therapeutic effects on chronic pain, as it does in post-stroke hemispatial neglect. Results of three studies have shown that PA performed with the affected hand to produce pointing after-effects towards the CRPS-affected side reduced pain and other CRPS symptoms [23–25]. One proposed mechanism of these apparent therapeutic effects is that PA reduces pain through correcting the lateralised spatial attention bias in people with CRPS. The magnitude of spatial biases has been previously linked to the severity of pain and other clinical signs of CRPS [7, 8, 26–30]. Moreover, PA leading to the after-effects away from the affected limb appears to increase pain in CRPS [25], further supporting the role of lateralised attention effects. Another potential mechanism is that PA restores normal sensory-motor integration. Although empirical evidence to support this mechanism is limited, it has been proposed that discrepancies between motor commands and sensory feedback can contribute to pathological pain, including CRPS [25, 31–33].

However, the studies demonstrating therapeutic effects of PA in CRPS [23–25] included only small numbers of patients (13 in total across all three studies), no sham treatment conditions, and were not blinded. Thus, to date there are no sufficient grounds for implementing PA as a standard rehabilitation method for CRPS [12]. The aim of this study is to provide a robust evaluation of the effects of PA on CRPS through a double-blind, randomized-controlled trial.

### Research questions and hypotheses

#### Primary research question (RQ) and hypothesis

RQ 1. Is two weeks of twice-daily PA treatment more effective in reducing pain and CRPS symptom severity than an identical regime using sham prism adaptation (“sham treatment”)?

Sham prism adaptation has an identical procedure to PA treatment, except that pointing movements are performed without any optical deviation and therefore no adaptation takes place. This will allow us to dissociate the effects of the additional movement of the affected limb imposed by the treatment, to isolate the true effects of PA.

> Hypothesis: There will be greater reductions in pain and CRPS symptom severity in the participants who receive PA treatment compared to the participants who receive sham treatment.

#### Secondary research questions and hypotheses

RQ 2. Are there any improvements in other clinical signs of CRPS, psychological functioning, and neuropsychological symptoms following PA treatment?

In addition to the primary outcome measures of pain and CRPS symptom severity, we aim to evaluate the effects of PA treatment on secondary outcomes (listed below) that are relevant to participants’ daily physical and psychological functioning, and for understanding the mechanisms of the therapeutic effects of PA (e.g., through establishing which neuropsychological symptoms might be affected by treatment).

> Hypothesis: Compared to the sham treatment group, participants in the PA group will have a reduction in spatial attention bias (consistent with its primary applications), as well as bias in cognitive representation of space and spatially-modulated motor function; body representation distortion (see [23]); emotional disturbance; fear of movement; average pain, movement restriction, and symptoms interference; and sensory, motor, and autonomic signs of CRPS following treatment.

RQ 3. How long are any benefits sustained for after the cessation of PA treatment?

We will determine this through assessment of all primary and secondary outcomes immediately and four weeks after the completion of treatment, and through additional assessment of a subset of self-reported secondary outcomes at three and six months post-treatment. The time course of any improvements will be also analysed at more granular level through participants’ daily subjective ratings of pain, range of movement, and the extent to which their symptoms interfere with daily life over a period of 10 weeks.

RQ 4. Are there factors that can predict the CRPS progression over time and / or the response to PA treatment?

Finally, the current study aims to explore potential predictors of the course of the disease and therapeutic response by tracking the symptoms of the same individuals over the course of 7.5 months. We plan to identify possible markers that would account for the individual differences in the progression of CRPS over time and / or in response to PA treatment. Due to insufficient evidence to support any specific predictions and limited sample size, we will perform exploratory analyses to address this research question. Factors such as demographic characteristics, pain intensity, CRPS symptom severity, sensory, motor and autonomic functions, and the extent of neuropsychological changes will be taken into consideration.

RQ 5. Are the neuropsychological changes in CRPS related to clinical signs and symptoms of CRPS?

A secondary aim of this study is to investigate the relationships between the severity of clinical symptoms of CRPS and the extent of neuropsychological changes in spatial attention, space and body representations, and motor functions.

> Hypothesis: Baseline abnormalities in perception of and attention to the affected limb and its surrounding space in participants with CRPS (compared to the perception and attention of healthy control participants) will correlate with the severity of pre-treatment clinical symptoms.

## Methods

### Design

This study has a double-blind, randomized, sham-controlled design. The schedule of enrolment, interventions, and assessments is presented in Table 1 and consists of four in-person Research Sessions (RS), two weeks of twice-daily home-based treatment, and two sets of long-term postal follow-up questionnaires. After provisional eligibility assessment through a structured phone interview, 42 participants with CRPS will undergo two baseline research sessions. Two baseline assessments (RS1 and RS2) are conducted to give an indication of normal fluctuations in CRPS symptoms (or lack thereof) prior to the treatment period. This will allow us to assess whether any change over the treatment period is meaningful, that is, greater than baseline fluctuations^1^. Research Session 1 (RS1) commences the timeline of the study at week 1 and includes in-person assessment of the eligibility criteria, informed consent, and collection of the outcome measures that are described in the “Measurements” section. Treatment allocation takes place 1-5 days before Research Session 2 (RS2), where the participants with CRPS are randomly allocated to one of the two groups of equal size: the PA treatment group or the sham treatment group. RS2 at the end of week 4 involves revisiting eligibility criteria and collecting outcome measurements. Immediately after completing RS2, the participants are instructed in how to carry out the treatment by a researcher who is not involved in any part of data collection. They then perform their first treatment under the guidance of that researcher. All other elements of the study (telephone screening, symptom assessment, experiment administration, and input of questionnaire data) are performed by researchers who are blind to the conditions that the participants have been allocated to. The treatment period spans weeks 5 and 6 of the study, where the participants perform twice-daily treatment in a self-guided manner. Outcome measurements are collected in two post-treatment assessments (RS3 and RS4) to evaluate differences in PA versus sham treatment effects, and whether any benefits of treatment are maintained at 4 weeks after treatment. The first post-treatment Research Session (RS3) takes place at the beginning of week 7 (i.e., the day immediately following the final treatment session). Research Session 4 (RS4) takes place in the beginning of week 11. Each research session is expected to last between 2 and 4 hours, including breaks between the assessments. During the first 10 weeks of the study, the participants also record their self-reported daily ratings of pain intensity, range of movement, and the extent to which their symptoms interfere with daily life in a provided logbook, which will allow us to track the time course of any changes between research sessions. Long Term Follow-Up 1 at 3 months (LTFU1; week 19) and Long Term Follow-Up 2 at 6 months (LTFU2; week 31) post-treatment are conducted through questionnaires sent and returned by post. RS3 marks the primary endpoint and LTFU2 marks the secondary and final endpoint of the study.

**Table 1.** Schedule of enrolment, interventions, and assessments for the participants with CRPS

| TIME POINT | STUDY PERIOD |  |  |  |  |  |  |  |  |
| --- | --- | --- | --- | --- | --- | --- | --- | --- | --- |
|  | Enrolment | Baseline research sessions |  | PA treatment or sham treatment (14 days) |  | Post-treatment research sessions |  | Follow-up |  |
|  | Prior to Week 1 (phone) | Week 1 RS1 | Week 4 RS2 | Week 5 | Week 6 | Week 7* RS3 | Week 11 RS4 | Week 19 LTFU1 | Week 31** LTFU2 |
| ENROLMENT: |  |  |  |  |  |  |  |  |  |
| Eligibility assessment | X | X | X |  |  |  |  |  |  |
| Informed consent |  | X |  |  |  |  |  |  |  |
| Allocation |  |  | X |  |  |  |  |  |  |
| INTERVENTIONS (one of two, twice-daily): |  |  |  |  |  |  |  |  |  |
| PA treatment OR |  |  |  | ◆ | ◆ |  |  |  |  |
| Sham treatment |  |  |  | ◆ | ◆ |  |  |  |  |
| ASSESSMENTS: |  |  |  |  |  |  |  |  |  |
| Self-report questionnaires |  | X | X |  |  | X | X | X | X |
| Clinical assessments |  | X | X |  |  | X | X |  |  |
| Computer-based tests |  | X | X |  |  | X | X |  |  |
| Daily logbook*** |  | ◆ |  |  |  |  | ◆ |  |  |

RS: Research Session; LTFU: Long Term Follow-Up (postal questionnaires only); PA: Prism Adaptation; \* Primary endpoint of the study.
\*\* Secondary endpoint of the study.
\*\*\* Self-reported average levels of pain, range of movement and symptoms interference with daily life in the last 24 hours, rated daily on 0-10 Numeric Rating Scales

Deviations from the schedule of consecutive Research Sessions and Follow-Ups will be accepted within the following time windows: up to 2-weeks deferral of RS2 and RS4, up to 1-week deferral of RS3, up to 3-weeks deferral of LTFU1 and LTFU2. If the times that the participant can attend RS2 and RS3 are planned to be longer than 14 days apart, the participant would commence the treatment 2 weeks before RS3. If the participant already started the treatment and has to postpone RS3, they would continue the treatment until RS3.

Twenty-one healthy control participants are being recruited for a single research session to obtain normative data. They undergo testing only once and do not receive any treatment.

#### Setting of the study

All research centres and recruitment sites are located in the United Kingdom. The University of Bath is the main research centre and one of the research sites, and research sessions can also take place at the Universities of Oxford, Exeter, or Liverpool; or in participants’ homes.

### Participants

#### Eligibility criteria

##### Participants with CRPS

This study enrols both male and female individuals, who:

1. are willing and able to give informed consent to take part in the trial,
2. are aged 18-80,
3. have a diagnosis of CRPS type I based on the Budapest diagnostic research criteria [3] as assessed at RS1 and revisited at RS2,
4. have CRPS type I primarily affecting one upper limb,
5. have had CRPS for a minimum of 3 months at the time of RS1,
6. and report current pain intensity ≥2 on a 0-10 Numeric Rating Scale at RS1 and RS2.

Participants are excluded from the CRPS group if they:

1. lack sufficient English language ability to provide informed consent,
2. are classified as legally blind,
3. have a history of neurological disorder (e.g. stroke, neurodegenerative disease or traumatic brain injury),
4. have CRPS meeting the Budapest diagnostic clinical or research criteria affecting both sides of the body^2^,
5. report that they have confirmed presence of nerve damage (CRPS type II) based on the results of nerve conduction test,
6. have dystonia or any other physical limitation severe enough to prevent satisfactory execution of PA / sham treatment,
7. or have a severe psychiatric comorbidity (such as schizophrenia) that in the researchers’ opinions would compromise participation in the study.

### Healthy control participants

The inclusion criteria for healthy control participants of this trial are:

1. willingness and ability to give informed consent,
2. age 18-80,
3. and being neurologically healthy and without current or chronic pain.

Criteria that would exclude an individual from the study are:

1. insufficient English language ability to provide informed consent,
2. being classified as legally blind,
3. physical disability or injury limiting normal mobility,
4. or a history of a neurological or severe psychiatric illness.

Each healthy control participant is matched to one participant with CRPS by sex, self-reported handedness prior to the onset of CRPS, and age (+/- 5 years).

#### Recruitment and participant retention strategies

The recruitment commenced on 31 March 2017 and is ongoing at the time of submission. People with CRPS are recruited through the National CRPS-UK Registry, Oxford University Hospitals NHS Foundation Trust, the Walton Centre NHS Foundation Trust, and other hospitals in the UK by post and clinicians’ referrals. Information about the trial is also disseminated through word of mouth, print and online advertisements and articles, and social media. Trial webpages have been set up on the funder’s and research centre’s websites. All of the above information channels provide potential participants with contact details of the authors, should they be interested in more information and / or taking part in the study.

To promote retention, participants are sent reminders before each RS and LTFU. Since recruitment takes place over a broad geographic area, their travel costs are reimbursed, or the research sessions are conducted in their own home. In recognition of the inconvenience of participation, which is heightened due to the burden of CRPS, participants receive a financial compensation of £250 for their time and contribution to the study once they complete RS4, and further financial compensation once they return the completed LTFU2 questionnaires by post (£50). Healthy control participants are reimbursed for their time and contribution at a rate of £10 per hour of their involvement.

Since the assessments and treatment are non-invasive and do not interfere with the participants’ ongoing standard treatment, and there are potential benefits from taking part, we expect good participant retention. Some participants may directly benefit from reduction in pain and CRPS symptom severity due to treatment. All participants will have an opportunity to undergo the PA treatment after the trial is completed, should the trial support the effectiveness of the treatment.

In the event of the participant’s withdrawal from the study, their data from any completed research sessions will be included in the analysis as far as possible. Participants who withdraw after RS2 will be considered lost to follow-up. For any participant who withdraws before RS4, an additional participant will be recruited to the trial such that there will be 42 full datasets for RS1-RS4. This strategy is implemented to assure sufficient and similar number of participants in each treatment arm. To address any potential selection bias, we will use intention to treat as our primary analysis, and per-protocol as supportive analysis (see “Treatment outcome analyses” section). Should participants deviate from the intervention protocol (e.g., missed treatment sessions), the number of logged treatment sessions can be used as a possible covariate in the final analyses.

### Randomisation

Treatment allocation is conducted by method of randomisation with stratification to minimise baseline (RS1) group differences. Eligible participants with CRPS are allocated in equal numbers to one of the two treatment groups: PA treatment group or sham treatment group. Group allocation is performed using MINIM computer programme [34] by a researcher who is not involved in data collection (JHB). The minimisation procedure controls for the stratification factors that are listed in Table 2. In the event of participant’s withdrawal after treatment allocation, but before RS3, their data shall be removed from the minimisation procedure and an additional participant shall be recruited for the trial to ensure equal numbers of full datasets with any post-intervention data in the two groups.

**Table 2.** Criteria for stratification as recorded in RS1

| Factor | Weight | Categories |
| --- | --- | --- |
| Current pain intensity (0 – 10 Numerical Rating Scale) | 2 | $\leq 6$<br>$> 6$ |
| CRPS severity score (1 - 16) [43] | 2 | $\leq 12$<br>$> 12$ |
| Primarily affected arm | 1 | Left<br>Right |
| Pre-CRPS dominant hand (writing) | 1 | Left<br>Right |
| Sex | 1 | Male<br>Female |
| Age | 1 | 18 - < 40<br>40 - < 61<br>61 - 80 |
| Presence of CRPS in body parts other than the primary affected arm | 1 | Yes<br>No |
| Presence of other non-CRPS pain | 1 | Yes<br>No |
| CRPS duration | 1 | < 1 year<br>1 - < 5 years<br>5 - < 10 years<br>$\geq 10$ years |

### Treatment

Participants in the PA treatment group are provided with welding goggles fitted with 35-diopter (Δ) Fresnel lenses that induce a visual shift of approximately 19° away from the CRPS-affected side. The optical displacement is of a similar magnitude as in previous CRPS studies that reported significant reductions in pain [23–25]. In contrast, no pain reduction was observed when a CRPS patient underwent two weeks of PA using lenses that shifted the visual image only by 5° [25]. Furthermore, prisms strength of 10°-15° was found to be sufficient to induce lasting amelioration of hemispatial neglect after brain injury [15, 18, 19, 35, 36], whereas weaker prisms did not improve neglect [37]. During each treatment session, the participant is seated in front of a vertical surface, such as a wall, upon which an A4 laminated page in landscape orientation is positioned. The page displays two visual targets (red circles 2cm in diameter), in each top corner. The page is mounted approximately at eye-level, hence targets are located 12.5cm (approximately 10°) to the left and to the right of the participant’s body midline. The distance between their torso and the wall is established individually, such that the participant can touch the targets with an almost fully extended arm (approximately 60cm). Participants put on the goggles and use their CRPS-affected arm to perform a total of 50 pointing movements (a number sufficient to induce sensory-motor adaptation [16]), alternating between the two targets (25 per side) and returning the pointing hand to their chest between each movement. The participants are instructed and trained to move as quickly as possible, and the goggles occlude the vision of the participant’s arm for approximately the first half of the movement. Both of these steps limit on-line correction of movement trajectory (strategic component of PA) and reinforce adaptive realignment, which is thought to maximise the effects of PA [13, 38, 39]. One treatment session takes approximately 5 minutes. The participant performs the treatment once under the guidance of an experimenter, and then twice daily for two weeks in a self-guided manner in their own home (giving a total of 29 treatment sessions). The intensity and duration of the treatment regime have been established based on previous studies evaluating the effects of PA on attention in hemispatial neglect following stroke, and on pain in CRPS. In particular, previous studies suggest that repeated sessions of PA are required to obtain a significant reduction in CRPS symptoms [23, 25] and that intense treatment (2 sessions a day for 4 days or more) produces symptom reduction that is sustained for at least two weeks post-treatment [23, 24].

Participants in the sham treatment group carry out the same procedure as the PA treatment group, except they are provided with goggles fitted with neutral lenses that do not induce optical deviation of the visual field. This is a standard control treatment for PA [18, 40]. Both prismatic and neutral lenses distort the acuity and clarity of vision, and both sets of goggles occlude the first part of the reaching movement. This factor ensures similarity of the two treatment arms in all aspects of the treatment aside from the sensory-motor adaptation.

To improve their adherence to the treatment protocol the participants receive in-person training, in which they complete the first treatment session guided by JHB or ADV, who ensure participants’ competence in performing the exercise according to the protocol. Furthermore, participants are provided with written instructions and a video tutorial. The researcher who trained them in the treatment is also available to address any questions or concerns about the procedure by phone or email. In order to monitor participants’ compliance and adherence, they keep a daily logbook throughout the treatment period, in which they record the time and duration of each treatment session. We will report the adherence to treatment as a percentage of participants in each treatment group who did not miss more than 6 treatment sessions. The extent of exposure in each group will be reported as average number of logged treatment sessions. Protocol deviations are defined as missed or additional treatment sessions, and sessions for which logbook entries suggest that anything other than the trained procedure has been used. We will report the total number of treatment sessions per group in which deviations other than missed or extra sessions are suspected. We will also compare the average number of logged treatment sessions between the two groups, and if significantly different, the number of logged treatment sessions will be used as a covariate in the analyses of the primary outcomes.

The participants are instructed to continue their standard pharmaceutical, physical, and / or other treatments during the trial, and are encouraged not to make any significant alterations to these treatments (e.g., major changes in medication, commencing new physiotherapy programmes). Medications and other treatments are noted during every research session to monitor any changes.

Criteria for discontinuing the allocated treatment before the 2 weeks have elapsed are a participant’s withdrawal from the study, or reports of experiencing an increase in CRPS symptoms that significantly heightens their discomfort or distress. As the treatment procedures require repeated movements of the CRPS-affected arm, participants may experience pain related to movement. However, this is expected to be temporary and no greater than the pain that could accompany standard physiotherapy or daily activities. To date, there have been no publications reporting serious adverse events related to PA in healthy controls or clinical populations (patients with stroke, Parkinson’s disease [40], or CRPS). In one case study exploring the effects of different PA directions and strengths, one CRPS patient experienced a small, temporary increase in pain when they performed PA using optical deviation towards the affected side [25]. Similar events in the current study are highly unlikely, as all PA is conducted with optical deviation away from the CRPS-affected side, i.e., in the direction thought to achieve therapeutic effects. Each participant is assigned their own dedicated set of prism goggles in a bag labelled with their participant code. The direction of optical deviation is independently checked by two people before the goggles are placed in a labelled bag. Any unexpected serious adverse events related to the administration of any study procedures will be reported to the researcher responsible for blinding (JHB) who will then make any decisions about discontinuing an individual’s participation and / or the trial, in consultation with the protocols for dealing with adverse events as outlined by the Research Ethics Committees.

### Measurements

Tests and measures used in the current study and time points at which they are administered are listed in Table 3. These are categorised as self-report questionnaires, clinical assessments, or computer-based tests.

**Table 3.** Measures

| Measurement domain | Measurement tool | Time points | Research question* / justification for use |
| --- | --- | --- | --- |
| **Self-report measures** |  |  |  |
| Pain and symptom interference | Current pain intensity (Item 6 of the Brief Pain Inventory) | Weeks 1, 4, 7, 11, 19 & 31 | RQ1, RQ3, RQ4, RQ5; group matching on baseline pain |
|  | Brief Pain Inventory† (BPI; short form; pain intensity and interference) [46] |  | RQ2, RQ3, RQ4, RQ5 |
|  | Pain Detect Questionnaire [49] |  | RQ2, RQ3, RQ4, RQ5 |
|  | Average pain intensity (Logbook) | Weeks 1 to 11 (daily) | RQ2, RQ3 |
|  | Average symptom interference (Logbook) |  | RQ2, RQ3 |
| Physical functioning | Average range of movement (Logbook) | Weeks 1 to 11 (daily) | RQ2, RQ3 |
|  | Edinburgh Handedness Inventory (current and change) [41] | Week 1 | RQ4, RQ5; participant characteristics |
| Body representation | Bath CRPS Body Perception Disturbance Scale (BPDS) [50] | Weeks 1, 4, 7, 11, 19 & 31 | RQ2, RQ3, RQ4, RQ5 |
| Emotional functioning | Tampa Scale for Kinesiophobia† [55] | Weeks 1, 4, 7, 11, 19 & 31 | RQ2, RQ3, RQ4, RQ5; group matching on baseline fear of movement |
|  | Profile of Mood States [51] |  | RQ2, RQ3, RQ4, RQ5; group matching on baseline mood disturbance |
|  | Revised Life Orientation Test [52] | Week 1 | RQ4; group matching on baseline levels of optimism |
| Treatment expectations | Patient-Centred Outcomes Questionnaire [53] | Week 1 | RQ4; group matching on expectations of treatment outcomes |
| Impression of treatment outcome | Patient's Global Impression of Change [54] | Weeks 7, 11, 19 & 31 | RQ2, RQ3 |
| Treatment adherence | Treatment sessions (Logbook) | Weeks 4 to 6 (twice-daily) | Monitoring treatment adherence |
| **Clinical assessments** |  |  |  |
| CRPS diagnosis | Budapest diagnostic research criteria assessment [66] | Weeks 1, 4, 7 & 11 | Verification of CRPS diagnosis and assessment of eligibility |
| Symptom severity | CRPS severity score [42] | Weeks 1, 4, 7 & 11 | RQ1, RQ3, RQ4, RQ5; group matching on baseline symptom severity |
| Autonomic functions | Limb temperature asymmetry | Weeks 1, 4, 7 & 11 | RQ2, RQ3, RQ4, RQ5 |
|  | Oedema |  |  |
| Motor functions | Grip strength | Weeks 1, 4, 7 & 11 | RQ2, RQ3, RQ4, RQ5 |
| | $\Delta$ Finger-To-Palm distance (ΔFTP) | | |
| Sensory functions | Mechanical Detection Threshold (MDT) | Weeks 1, 4, 7 & 11 | RQ2, RQ3, RQ4, RQ5 |
|  | Mechanical Pain Threshold (MPT) |  |  |
|  | Mechanical Allodynia |  |  |
|  | Two-Point Discrimination [75] |  |  |
| **Computer-based tests of neuropsychological changes** |  |  |  |
| Visuospatial attention | Visual Temporal Order Judgement (TOJ) [9] | Weeks 1, 4, 7 & 11 | RQ2, RQ3, RQ4, RQ5 |
|  | Landmark task [76] |  |  |
|  | Greyscales task [77] |  |  |
| Mental representation of space | Mental Number Line Bisection task [79] | Weeks 1, 4, 7 & 11 | RQ2, RQ3, RQ4, RQ5 |
| Spatially-defined motor function | Directional Hypokinesia [103] | Weeks 1, 4, 7 & 11 | RQ2, RQ3, RQ4, RQ5 |
| Body representation | Hand Laterality Recognition task [80] | Weeks 1, 4, 7 & 11 | RQ2, RQ3, RQ4, RQ5 |

\* RQ1, effects of treatment on the primary outcome measures; RQ2, effects of treatment on the secondary outcome measures; RQ3, time course / duration of any changes; RQ4, predictors of CRPS progression over time and/or response to treatment; RQ5, baseline abnormalities in neuropsychological functions in participants with CRPS compared to pain-free controls, and their relationships with clinical signs of CRPS (only Week 1 data).
† Brief Pain Inventory interference subscale and Tampa Scale for Kinesiophobia are also considered proxy measures of physical functioning.

#### Baseline descriptors

Age, sex, and handedness of all the participants are recorded as demographic characteristics. An interview regarding their medical history is conducted to collect information about the date and type of any inciting injury or insult, CRPS duration in months from diagnosis to RS1, the presence of CRPS in body parts other than the primarily affected upper limb, the presence of non-CRPS pain conditions and other co-morbidities, and current treatments.

A hand laterality index is calculated using the Edinburgh Handedness Inventory [41] in RS1. The scoring can range from -100 (extreme left-handedness) to 100 (extreme right-handedness). All participants respond regarding their current hand preference, and the participants with CRPS additionally complete another version of the Edinburgh Handedness Inventory based on their recalled hand preference prior to the onset of CRPS symptoms. A “change in handedness” score (Handedness after CRPS – Handedness before CRPS) is calculated to give an approximation of the functional impact of the CRPS.

#### Primary outcomes

A change between RS2 (immediately before the commencement of treatment) and RS3 (immediately after the end of the treatment period) in current self-reported pain intensity and CRPS severity score [42, 43] are the primary outcomes. People with CRPS consider pain relief to be the highest priority for recovery [44], and pain intensity is the most common primary outcome in chronic pain trials [45]. Current pain intensity is measured using item 6 of the Brief Pain Inventory (BPI; short form) [46], which is a Numerical Rating Scale (NRS) ranging from 0 – “no pain” to 10 – “pain as bad as you can imagine”. The BPI has high reliability [46]. In addition to pain, CRPS involves a range of other debilitating symptoms, some of which were also affected by PA in previous studies [23, 25]. Therefore, we included a comprehensive measure of symptoms severity as the second primary outcome. The CRPS severity score assessment protocol follows the 16-points scoring system published by Harden and colleagues [43]. This continuous index of CRPS symptom severity has good discrimination abilities, concurrent validity and adequate sensitivity to change [42, 43], and has been recommended as one of the core outcome measures for CRPS clinical studies [47].

#### Secondary outcomes

##### Self-report questionnaires

There is a lack of validated outcome measures for CRPS (however, see recently published recommendations [48]). Therefore, the choice of the measures for the current trial was guided by general recommendations of core outcome measures for chronic pain clinical trials (IMMPACT; [45]) and the existing literature on CRPS implicating other relevant questionnaires.

There are 10 self-report questionnaire measures of pain, physical and emotional functioning, body representation, expectations about treatment, and impressions of treatment outcome. The BPI [46], Pain Detect Questionnaire [49], Bath CRPS Body Perception Disturbance Scale (BPDS; [50]), Tampa Scale for Kinesiophobia, and Profile of Mood States [51] are completed in every research session and long-term follow-up (RS1-RS4, LTFU1-LTFU2). A Revised Life Orientation Test [52] and a Patient-Centred Outcomes Questionnaire [53] are administered only in RS1. The Patient’s Global Impression of Change questionnaire [54] is completed only at post-treatment research sessions and long-term follow-ups (RS3-RS4, LTFU1-LTFU2). Finally, a daily logbook of self-reported average pain, range of movement, and symptom interference is kept by the participants during the baseline, treatment, and post-treatment periods (i.e., every day for the 10 weeks that elapse between RS1 and RS4).

Participants use the short-form of the BPI [46] to rate their pain intensity (current, average, and worst and least pain over the last 24 hours) and the extent to which pain interferes with their physical, social and psychological functioning on 0-10 NRSs (0 – “no pain” or “does not interfere”; 10 – “pain as bad as you can imagine” or “completely interferes”, respectively). The pain intensity component of BPI can result in an average score between 0 and 10; an average interference component score can also range from 0 to 10. The Pain Detect Questionnaire is a validated measure of the neuropathic features of experienced pain [49] scored from -1 to 38, with higher scores indicating a greater neuropathic component of pain.

The BPDS [50] includes seven self-reported items to assess subjective detachment, awareness, attention to, and feelings about the CRPS-affected limb; the perceived changes in size, temperature, pressure, and weight of the limb; and any desire to amputate the limb. The BPDS includes a mental imagery task in which the mental representation of both limbs (affected and unaffected) is sketched by a researcher based on the participants’ description. Total score ranges from 0 (no disturbance) to 57 (most severe disturbance of body perception). Since BPDS is not a validated measure, normative data is also collected from healthy control participants who are responding to the self-report components regarding the limb that corresponds to the CRPS-affected limb of their matched participant with CRPS.

The Tampa Scale for Kinesiophobia [55] is administered to measure pain-related fear of movement and re-injury. The participants choose the extent to which they agree with each of 17 statements about fear of movement and physical activity that could (subjectively) cause pain and / or injury (1 – “strongly disagree”, 4 – “strongly agree”). The final score varies from 17 to 68 points, with higher numbers indicating more severe kinesiophobia. The Tampa Scale for Kinesiophobia is included as a measure of the likely extent to which participants use their affected limb and their beliefs and emotions about those movements.

Considering that mood can exert effects on pain [56–58] and attention [59–61], the Profile of Mood States is administered in the current trial to verify that the two treatment groups are matched according to mood disturbance, and to enable evaluation of whether treatment results in any significant differences in mood improvements between the groups. The Profile of Mood States is a 64-item scale indicating the extent to which the respondent is experiencing various transient, distinct mood states (1 – “not at all”, 5 – “extremely”). High reliability and validity of Profile of Mood States [51, 62] has been reported. This measure is also completed by healthy control participants at a single research session.

The Revised Life Orientation Test [52] assesses levels of optimism and pessimism. Participants rate to what extent they agree with 10 statements on a scale from 0 – “strongly disagree” to 4 – “strongly agree”. The Patient Centred Outcomes Questionnaire [53] is also administered to measure patient-centred expectations and criteria for success in chronic pain treatment. Rating scales from 0 to 10 are used to indicate the usual, desired, expected and considered successful levels of pain, fatigue, emotional distress, and interference with daily activities (0 – “none”, 10 – “worst imaginable”), and the importance of improvement in each of these areas (0 – “not at all important”, 10 – “most important”). The decision to include the Revised Life Orientation Test and the Patient Centred Outcomes Questionnaire in the current trial was driven by the fact that optimism and expectations of outcome have been known to influence the success of novel treatments [63–65]. Thus, it is important to confirm that the two treatment groups are matched on these extraneous factors, or to include these variables as covariates in the analysis of outcome measures if they are not.

The participants keep daily logbooks for weeks 1-11 in which they use 0-10 NRSs to record their average level (over the preceding 24 hours) of pain (0 – “no pain at all”, 10 – “pain as bad as it could be”), the range of movement in the affected arm (0 – “no movement at all”, 10 – “normal movement”), and the degree to which their symptoms have interfered with their daily life (0 – “no interference at all”, 10 – “complete interference”). These measures are designed to track the time-course of any change in pain, movement, and interference during the first 10 weeks of the study (i.e. four-week baseline period, two-week treatment period, and four-week immediate post-treatment period).

Finally, the Patient Global Impression of Change questionnaire [54] is administered to measure participants’ impression of how much their symptoms have changed due to treatment. It produces a single rating on a scale from 1 – “no change” to 7 – “a great deal better”. The Patient Global Impression of Change is a widely recommended measure of perceived global improvement and satisfaction with treatment [45, 48].

##### Clinical assessments

The clinical measures include examination of CRPS signs and symptoms, sensory thresholds, autonomic changes, and motor functions. Participants with CRPS undergo all clinical assessments in RS1-RS4, whereas healthy control participants undergo the same clinical assessments during a single research session. Locations for sensory testing are the most painful site on the CRPS-affected limb and the corresponding site on the unaffected limb, always beginning with the unaffected limb so that participants can be familiarised with the test procedures and sensations before the tests are administered on their painful limb. For sensory testing in control participants, measures taken from the limb corresponding to the CRPS-affected limb of their matched participant with CRPS are compared to measures taken from the other limb.

CRPS diagnosis is confirmed in RS1 and RS2 during the baseline period, before commencement of the treatment, based on the Budapest research criteria [66]. These criteria are also assessed in the post-treatment period (RS3-RS4) to determine if the participants still meet the CRPS diagnosis.

The severity of symptoms is assessed and quantified as CRPS severity score in RS1-RS4, according to a recently validated protocol [42, 43]. Each of the 16 items is recorded as present (“1”) or absent (“0”) based on the self-reported symptoms and the signs confirmed at the time of examination through sensory testing, visual, and manual assessments. These include continuing, disproportionate pain; allodynia; hyperalgesia and / or hypoesthesia; temperature, colour, and sweating asymmetry; oedema; dystrophic changes; and motor abnormalities. Summed scores indicate the overall CRPS severity score. Where possible, criteria are evaluated based on a comparison between the affected and unaffected upper limb for a sign to be classified as present, including objective quantification of limb temperature asymmetry, oedema, muscle weakness, and active range of movement.

Photographs of the dorsal and palmar surface of both hands and forearms are taken so that the presence of skin colour and trophic changes can be double-scored by a researcher who is not involved in data collection and who is blind to the time point at which the photographs were taken, to which limb is affected by CRPS, and to which group the participant is allocated. Video recordings of both limbs performing the movements of fist closure and opening, wrist flexion and extension, and radial and ulnar wrist deviation are taken so that the motor abnormalities can be double-scored according to the same protocol. We will use Cohen’s kappa statistic to report inter-rater agreement.

An infrared thermometer is used to measure temperature asymmetry. Temperature measurements are taken to the nearest 0.1°C from the dorsal and palmar surface of both hands (over the thenar muscle) and the centre of the region of worst pain as indicated by the participant. An arithmetic mean of the 3 measurements on each limb is calculated. According to the Budapest diagnostic criteria [66], an absolute difference between the affected and unaffected side greater than 1°C is classed as a temperature asymmetry. When available, thermal images of both limbs are additionally taken (camera FLIR T620 that is sensitive to changes in temperature as small as 0.04°C).

Oedema is measured using the figure-of-eight procedure that uses a soft tape measure. The detailed protocol for hand and wrist size measurement is described elsewhere [67]. This measure has excellent intra- and interrater reliability and concurrent validity compared with water volumetry [68]. Hand size is calculated as an arithmetic mean of 3 measurements performed on each hand. Presence of asymmetric oedema is considered if the average measure taken from the CRPS-affected hand is at least 0.56cm larger compared to the unaffected hand, which was suggested to be a clinically relevant difference in a previous study [69].

Grip strength is measured as a marker of muscle weakness, using an electronic hand dynamometer (Constant, model 14192-709E). Participants are seated in a chair with their elbows flexed at 90°, forearms in neutral position, and wrists at between 0 and 30° extension. They are instructed to squeeze the dynamometer’s handle as hard as they can and perform three such trials with each hand, alternating between the hands and allowing a pause of at least 15 seconds between each trial. An arithmetic mean of the 3 measurements (kg force) for each hand is calculated. Muscle weakness of the affected hand is indicated if the ratio of grip force in the affected to unaffected side is smaller than 0.95 for left-handed participants or smaller than 0.85 for right-handed participants. These criteria take into account the normal difference between dominant and non-dominant hands for left- and right-handed individuals [70, 71].

Active range of movement in the hands is assessed by measuring a change in Finger-To-Palm (ΔFTP) distance (cm). A detailed measurement protocol is described elsewhere [72]. ΔFTP is an index of the extent to which a person can fully flex their fingers (e.g., to make a fist) relative to the extent to which they can extend them (e.g., to make their hand flat). ΔFTP was selected as a measure of range of movement as it takes into account both these aspects of motor function, unlike classic FTP that only regards the maximum flexion. A significant decrease in the range of movement in the affected hand is defined as ΔFTP_affected_ / ΔFTP_unaffected_ < 0.9.

In addition to those limb differences that are assessed through clinical examination for the CRPS severity score, differences between the affected and unaffected limbs are also objectively quantified through elements of a standard Quantitative Sensory Testing (QST) procedure to assess hypoesthesia, pinprick hyperalgesia, and allodynia. Participants undergo the assessment of Mechanical Detection Threshold (MDT) that follows the standardized protocol [73] using von Frey filaments of 0.008g to 300g force (Bioseb, model Bio-VF-M). Then the ratio of thresholds for affected vs. unaffected side is derived [(MDT_affected_-MDT_unaffected_)/MDT_affected_]. A positive score indicates hypoesthesia (increased tactile detection threshold) on the affected side. Based on relative QST reference data comparing both sides of the body, hypoesthesia is confirmed if the ratio is ≥ 0.38 [74]. We also assess Mechanical Pain Threshold (MPT) according to the standardized protocol [73] on both limbs, using pinprick stimulators of 8mN to 512mN intensities (MRC Systems PinPrick Stimulator Set). A positive thresholds ratio [(MPT_unaffected_-MPT_affected_)/MPT_unaffected_] indicates hyperalgesia (decreased pain threshold) on the affected side. Hyperalgesia is confirmed if the ratio is ≥ 0.4, based on relative QST reference data comparing both sides of the body [74]. Allodynia is examined using a procedure adapted from the dynamical mechanical allodynia test of the QST [73]: the cotton ball, Q-tip and brush (MRC Systems PinPrick Stimulator Set) are applied to the skin five times each, in a random order, with a single 1-2cm long sweeping motion lasting approximately 1 second. Participants rate each sensation on a scale from 0 – “no pain, no sharp, pricking, stinging, or burning sensation” to 100 – “most intense pain sensation imaginable”. Any sharp, pricking, stinging, or burning sensation is defined as painful and given a rating above 0. Allodynia is quantified as an arithmetic mean of 15 ratings on each limb. Its presence is indicated by scores greater than zero.

A Two-Point Discriminator disk (Exacta, North Coast Medical) is used to record tactile discrimination thresholds [75]. The participant’s index finger tip is touched either with one tip or two tips of the disk for 3 seconds per touch, with consistent pressure, and while the participant has their eyes closed. On each trial, participant reports whether they perceived touch on one point or two points of their finger. The procedure starts with two points separated by 7mm distance, and the distance between points is increased or decreased (down to a single tip) across trials according to the staircase procedure. For example, if the participant initially reports two touches, smaller distances are applied until the participant reports the sensation of only one point. The distance is then increased until a sensation on two points is reported again. The procedure continues until 5 subthreshold and 5 suprathreshold values are obtained. The tactile discrimination index is calculated as a geometric mean of these 10 turning points for each hand. To quantify the difference between the two sides of the body, we derive the tactile discrimination thresholds ratio [(affected-unaffected)/affected]. Positive score indicates less precise tactile discrimination ability on the affected limb.

##### Computer-based / psychophysical tests

Six computer-based measures are used in the present study to assess the following neuropsychological functions: visuospatial attention, cognitive representation of space, spatially-defined motor function, and body representation. To test for spatial attention bias in near space, we administer versions of three tasks that have been used to measure spatial attention in hemispatial neglect: a visual Temporal Order Judgement (TOJ) task [9], a Landmark task [76], and a Greyscales task [77]. The fourth task is a Mental Number Line Bisection task, which measures the mental representation of space [78, 79]. The fifth task is a Directional Hypokinesia task, a measure of motor “neglect-like” impairment. The final computer-based task is a Hand Laterality Recognition task, which is thought to be indicative of body representation [80].

All measures presented in this section are collected in RS1-RS4 from the participants with CRPS and during a single research session from healthy control participants. Hand and side of space for all tasks are coded as affected or unaffected (for controls, the “affected” and “unaffected” hand / side is coded based on their matched participant with CRPS). Each task is preceded by a short practice session to familiarise the participant with the task. If they do not appear to follow the instructions during practice, these are explained again, and the practice is repeated.

##### Visuospatial attention

The following three computer-based tests are used to measure visuospatial attention: the visual TOJ task, the Landmark task and the Greyscales task.

#### The visual TOJ task

TOJ tasks are sensitive measures of covert spatial attention, used both in clinical populations [81–87] and healthy people [88–92]. The usual procedure involves presenting pairs of identical stimuli, one on each side of space, with different onsets but the same duration. The participant’s task is to report which of the two stimuli they perceived first. According to the prior entry hypothesis [93], stimuli that are subject to greater attention are perceived earlier relative to stimuli that are subject to lesser attention. The TOJ task takes advantage of this premise. The visual variant of the TOJ used in this study is similar to that described in a previous article [9]. The participants keep their hands uncrossed on their laps under the table, and have their head stabilised by a chinrest. They are instructed to maintain their gaze on a black fixation point (3mm in diameter), approximately 28cm from their torso, located in the centre of a 46.5 × 35.5cm white board laid on a table. Pairs of brief (10ms) red light stimuli (3mm in diameter) are presented using laser pointers controlled via an Arduino platform that is integrated with PsychoPy software [94]. The lights are presented 9cm (approximately 18°) to the left and 9cm to the right of the fixation point (one on each side), using a range of ten temporal offsets: ±10, ±30, ±60, ±120 and ±240ms (with negative numbers representing the trials in which the light on the affected side appeared first). Each temporal offset is presented 15 times in pseudorandom order, giving a total of 150 trials. To account for any response bias [95] the participants complete the TOJ task once while indicating which of the two lights appeared first, and a second time while indicating which light appeared second (order counterbalanced between participants). Participants’ verbal responses (“Left” or “Right”) are inputted via the computer keyboard by the researcher. The relative number of left-right responses to different offsets of the stimuli is re-expressed as the number of affected-unaffected responses. To derive the Point of Subjective Simultaneity (PSS) for each participant and each condition, these data are then fitted with a cumulative Gaussian using a criterion of maximum likelihood. The PSS expresses the amount of time (ms) by which the light that appears in the affected side of space should precede (negative PSS values) or follow (positive PSS values) the light that appears in the unaffected side of space for the two stimuli to be perceived as simultaneous. For the analysis, PSSs from the two response blocks (which light appeared first or second) will be averaged to obtain a single index of attention bias. A negative PSS value indicates a bias of attention away from the affected side, whereas a PSS value of 0 indicates equal distribution of attention to both sides of space.

#### The Landmark task

In addition to the TOJ task, participants complete four tasks involving presentation of visual stimuli on a computer screen. For these, participants are seated with their head in a chinrest that is aligned with the centre of the screen. Stimuli are presented on a laptop touch screen (34.5cm × 19.4cm size, 1920 × 1080 pixels resolution) using PsychoPy software [94] on the Windows 10 operating system. The laptop screen is positioned at a viewing distance of 50cm. The responses are recorded using a custom-made button box positioned such that the buttons are aligned vertically.

We use a modified version of a Landmark task to measure bias in attention to or the representation of relative horizontal distance in near space. The task is adapted from a previous study [76] and involves simultaneous presentation of two stimuli (“landmarks”; white circles 1.1° in diameter) to the left and to the right of a central fixation cross. The total distance between the two landmarks is kept constant across trials (15°), however, their position relative to the fixation cross varies by 0.1° increments from ±8.1° to ±6.9° away from the fixation cross in the horizontal plane (Figure 1). Thus, there are 6 stimulus pairs in which the right landmark is closer to the fixation cross, 6 stimulus pairs in which the left landmark is closer, and 1 stimulus pair in which the distance of both landmarks from the fixation cross is equal. Each stimulus pair is presented 15 times during one block resulting in 195 trials per block, presented in pseudorandom order. The participant is instructed to maintain their gaze on a white, 1.4° high fixation cross presented in the centre of a grey screen. After 500ms, the fixation cross is joined by the two stimuli which are displayed for 300ms. Then a 200ms mask is presented, consisting of a white 1.6° high line extending horizontally across the entire screen, with a grey fixation cross in the same location as the previous white one (Figure 2).

**Figure 1.**
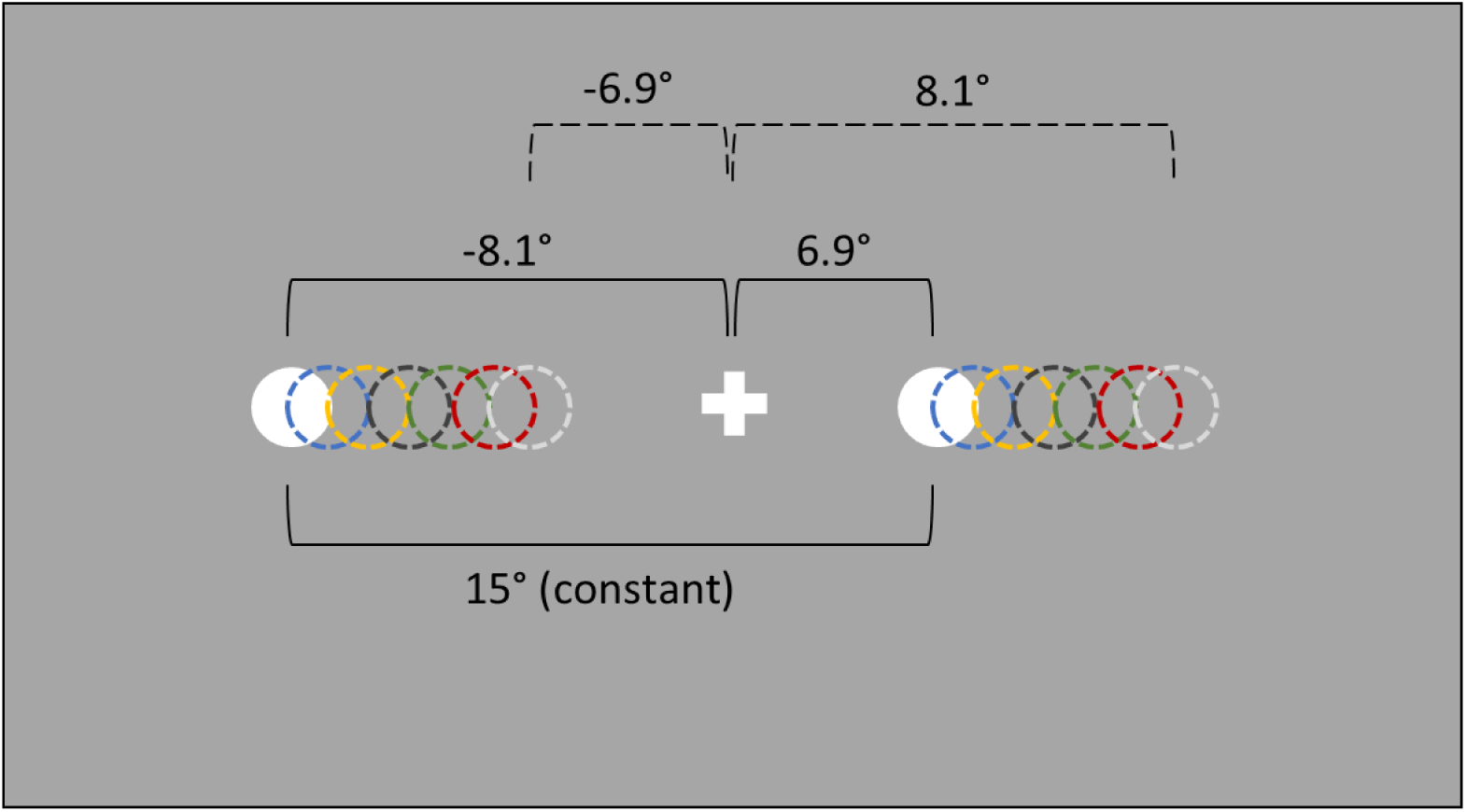
Representation of the stimuli in the Landmark task. White filled circles represent the stimulus pair in which the left landmark is farther from the fixation cross (-8.1° away) and the left landmark is closer (6.9° away). Circles with dashed lines in matched colours represent other possible stimulus pair locations.

**Figure 2.**
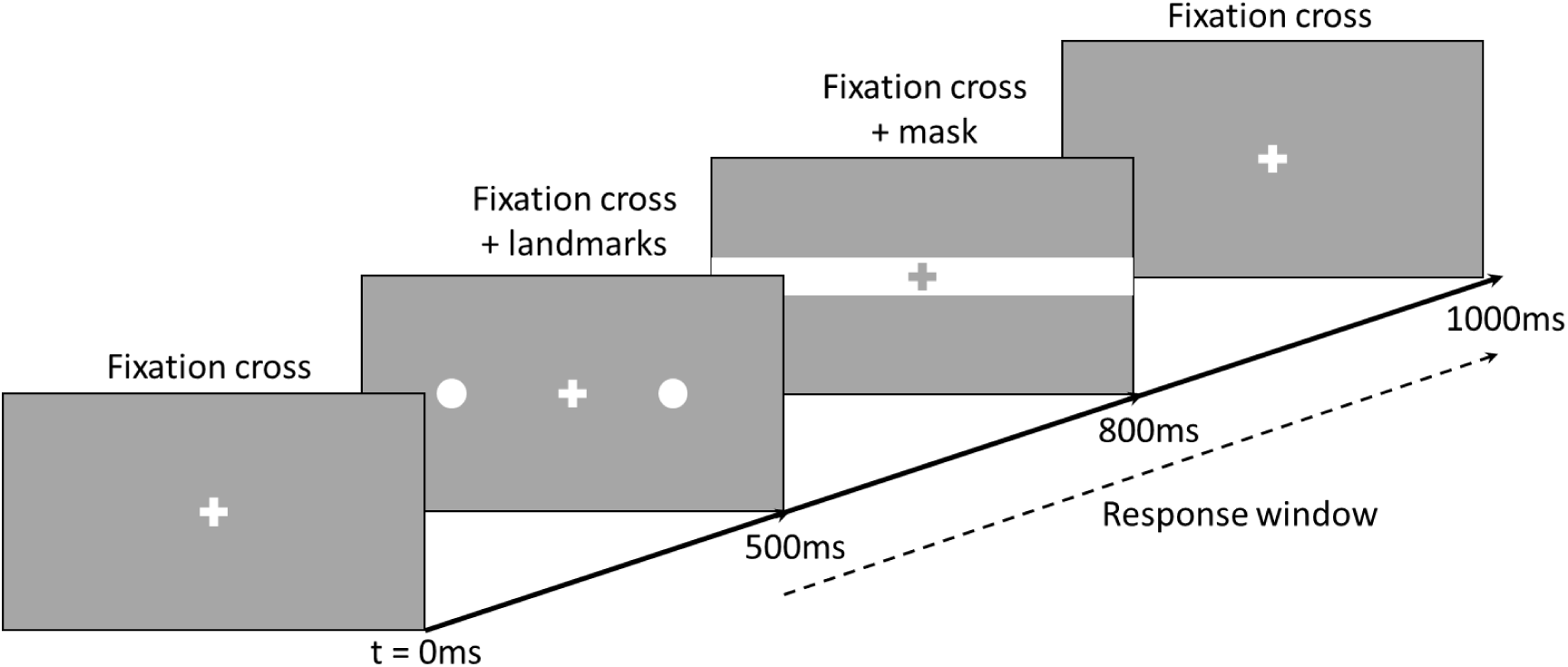
The time course of a single trial in the Landmark task.

Participants are instructed to indicate whether the left or the right landmark was closer to the fixation cross. They give their responses by pressing the green (“left”) or red (“right”) button (using the index and middle finger of the unaffected hand). The button press ends the trial and initiates the next trial. To control for response bias, in a separate, second block of the task, they are instructed to indicate which target was further away from the fixation cross by pressing the same buttons. The order of the two blocks is counterbalanced between participants. Attention bias is calculated from a relative number of “Left” and “Right” responses to each stimulus pair (landmarks position relative to the fixation cross). This is re-expressed in terms of affected versus unaffected sides of space and converted to a Point of Subjective Equality (PSE) using a cumulative Gaussian fit. The PSE expresses the relative distance at which the landmark on the affected side of space should be further from (negative PSE values) or closer to (positive PSE values) the fixation cross for the two landmarks to be perceived as appearing at equal distance from the fixation cross. A negative PSE value indicates an attention bias away from the affected side and / or under-representation of that side of space. For example, if a participant with a left-affected limb indicates that the left landmark is appearing closer to the fixation cross more often than the right landmark (i.e., underestimating distance on left side), their PSE value will be negative and indicate reduced attention to or under-representation of the left (affected) side. We will average the PSEs from the two response blocks (which landmark was closer or further away from the fixation cross) to obtain a single spatial bias index for our analyses.

#### The Greyscales task

The Greyscales task is a sensitive measure of overt spatial attention bias. The task used in the present study follows a previously developed procedure [77]. Forty pairs of short (9.95° × 1.95°) and long (12° × 1.95°) greyscale bars (Figure 3) are presented in the centre of a white screen in a free-viewing condition until the response is given. Participants indicate if the top or the bottom bar appears overall darker by pressing the upper or lower button, respectively (using the index and middle fingers of their unaffected hand). The trials are separated by an 18° × 8° mask (random dot 1111 × 362 black and white pixel pattern of static) displayed for 150ms, after which the next trial begins. An attention bias score is calculated by subtracting the number of “rightward” responses (choosing whichever bar is darker on its right side, regardless of its vertical position) from the number of “leftward” responses and dividing the difference by a total number of trials. Negative scores indicate rightward bias, i.e. reduced attention to the left side. This will be re-expressed as bias relative to the affected / unaffected side.

**Figure 3.**
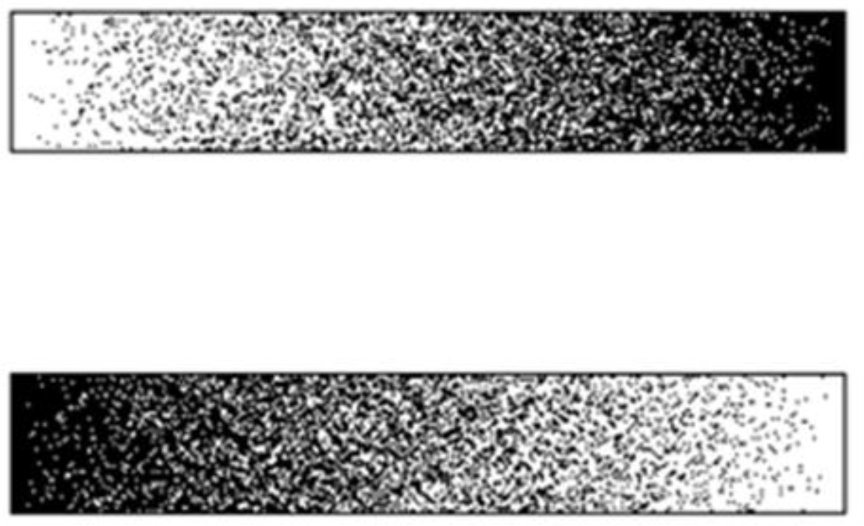
Example pair of stimuli in the Greyscales task. A person who has reduced attention to the left side of space would judge the upper bar as having overall greater average darkness.

##### Mental representation of space

The Mental Number Line Bisection task aims to measure spatial bias in the mental representation of space. This is based on the evidence that people implicitly represent numbers in a linear arrangement in which smaller numbers are located to the left side of space, and larger numbers are located to the right side of space [96]. The procedure is adapted from a previous study [79] in which pairs of numbers were read aloud to the participants and they were required to indicate the number that would fall midway between the two without making any calculations. The current task uses the same intervals (9, 16, 25, 36, 49 and 64) between two numbers that ranged from 2 to 98. For example, the midpoint number between 54 and 70 (16-interval) would be 62 (Figure 4). The only deviation from the previous procedure [79] is that every pair of numbers is presented twice – once in ascending and once in descending order, to reduce response bias. There are 84 trials presented in pseudorandom order and participants’ verbal responses are inputted to the computer via the keyboard by the researcher. We subtract the subjective midpoint number from the objective midpoint number in each trial (for example, see Figure 4), and the average score is transformed to indicate the relative bias in the mental representation of space away (negative values) or towards (positive values) the affected hand-side. A bias away from the affected side was previously found in CRPS patients on Mental Number Line Bisection [79], as well as a rightward bias in post-stroke hemispatial (left) neglect patients [78, 97–99], and a leftward bias in healthy participants (“pseudoneglect”) [79, 100, 101].

**Figure 4.**
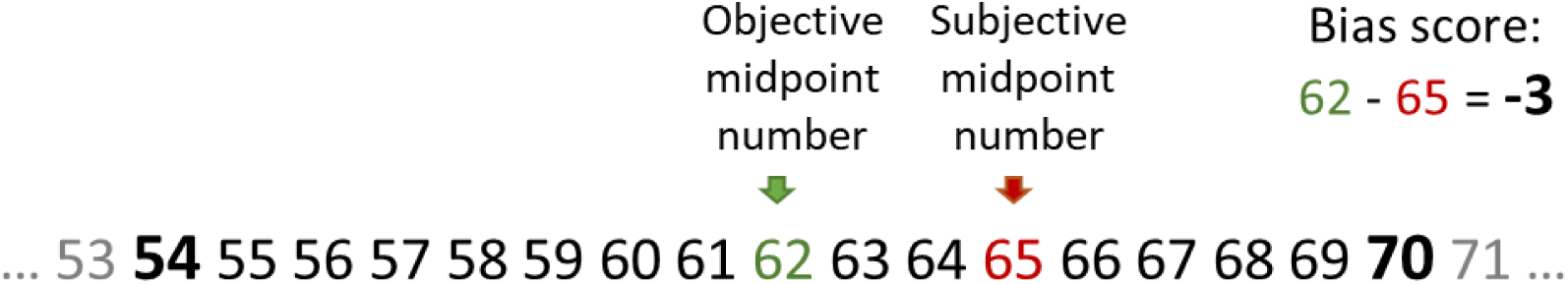
A pictorial representation of a theoretical trial from the Mental Number Line Bisection task. The participant is asked to indicate the midpoint number between the numbers 54 and 70, which are verbally presented by the experimenter. A negative bias score indicates that the centre of the participant’s mental number line is shifted towards larger numbers, consistent with an under-representation of the left side of space relative to the right side of space.

##### Spatially-defined motor function

We use the Directional Hypokinesia task to assess two distinct forms of motor neglect – directional hypokinesia, i.e. relative slowing in the initiation of movements directed toward the affected side, and directional bradykinesia, i.e. relative slowing in the execution of movements directed toward the affected side of space [102]. The task measures movement initiation and execution times to targets that appear on the left or the right side of the screen. The task follows the exact procedure previously used for research with hemispatial neglect patients [103]. A black 1.4° fixation cross and two black 3° × 3° squares, one 12° to the left and one 12° to the right of the fixation cross, are on constant display (locations are re-expressed as affected and unaffected Visual Field, VF). Each trial is initiated by the participant pressing and holding a button with their index finger. After a time interval that varies randomly between 1500ms and 3000ms a black target (1.4° high “X”) appears inside one of the squares, in a pseudorandomized order, for 2000ms. The target onset initiates the response window and the participant is required to release the button, touch the screen in the location where the target appeared, and then return their index finger to the button as fast as possible, which initiates the next trial. There are 30 trials per block. A touch screen is used to monitor the accuracy of pointing-to-target movements. The Reaction Times (RTs) to release the button after the target onset (Movement Initiation Time, MIT) are recorded, as well as time taken between releasing the button and touching the screen (Movement Execution Time, MET). There are three different hand Starting Positions (location of the button box): 25cm to the left from body midline, central (aligned with the body midline), and 25cm to the right from body midline (the locations are re-expressed as the affected, central, and unaffected side). Manipulating the hand Starting Position allows dissociation between perceptual component of the task (e.g., slower detection of the targets on the affected side) and the true directional hypokinesia. Participants perform each condition once with each hand in separate blocks, giving a total of 6 conditions (unaffected hand from the unaffected side, unaffected hand from the centre, unaffected hand from the affected side, affected hand from the unaffected side, affected hand from the centre, affected hand from the affected side). The order of the Starting Positions is counterbalanced between participants, with the only restriction that they alternate between the unaffected and the affected hand in each subsequent block to reduce fatigue.

We will calculate mean MITs and METs for each combination of VF in which the target appeared (affected and unaffected) and hand Starting Position (affected, central, and unaffected location), separately for each hand used to complete the task. Directional hypokinesia would be indicated by slower initiation of movements (MIT) towards the affected side of space, independent of which arm is used [102–104]. Directional bradykinesia would be indicated by slower movement execution times (MET) towards targets appearing in their affected side of space, even when using the unaffected arm.

To dissociate any signs of directional hypokinesia from potential mechanical constraints, two indices of directional hypokinesia will be derived based on the analyses described in previous research [103]. Movement pathways and indices are illustrated in Figure 5. The first index (A) quantifies the difference in MITs to the targets in the affected vs. unaffected VF as a function of the direction of the movements [i.e. reaching toward the affected side (from central Starting Position) relative to reaching toward the unaffected side (from affected Starting Position). Index A will be calculated as: [central Starting Position (MIT affected VF – MIT unaffected VF) – affected Starting Position (MIT affected VF – MIT unaffected VF)]. Thus, a larger value on this index will indicate greater directional hypokinesia. A potential drawback of Index A is that it involves planning a movement across body midline (from the affected Starting Position to the unaffected VF) that covers a longer distance and may be more difficult than other movement trajectories. Therefore, we will also derive a second index (B) that directly quantifies the relative slowing in the ability to initiate movements to the targets in the affected VF when making movements of the same physical length toward the affected side (from central Starting Position), versus toward the unaffected side (from affected Starting Position). Index B will be calculated as [central Starting Position (MIT affected VF) – affected Starting Position (MIT affected VF)]. Positive values on each index (A and B) would indicate hypokinesia for the affected side. Analogous indices A and B will be calculated for METs, and positive values of each index would indicate directional bradykinesia for the affected side.

**Figure 5.**
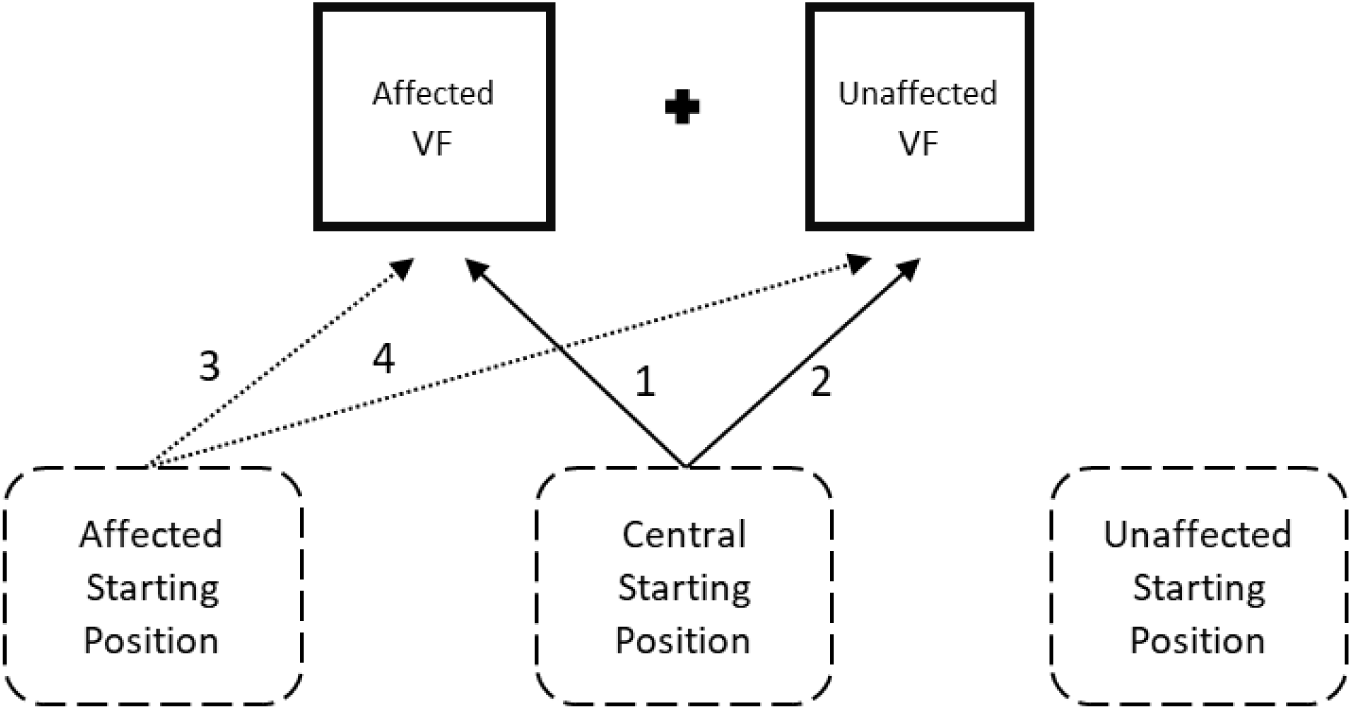
Indices of Directional Hypokinesia task. Target locations (affected and unaffected Visual Field, VF) and hand Starting Positions (affected, central, and unaffected) are presented as an example of a participant with CRPS of left arm. Index A is calculated as initiation time of the movements represented by arrows [(1 – 2) – (3 – 4)]. Index B is calculated as initiation time of movements (1 – 3).

##### Body representation

As an objective measure of body representation we use a modified Hand Laterality Recognition task based on a procedure described elsewhere [80]. The stimulus set was developed specifically for the current study (examples shown in Figure 6) and the final images were chosen based on the results of a pilot study reported in Additional file 2. The images depict gender-neutral right and left (mirror-reversed) hands in different postures and are presented at four different orientations (0°, 90°, 180° and 270°). In each trail, a black 0.1° fixation cross on a white background is on constant display. After 1000ms a colour image of a hand (12.9° × 12.9°) is randomly presented 8° to the left or to the right of the fixation cross (i.e., in the left or the right VF, as in a previously published similar procedure [8]) for 180ms. This period is short enough to prevent foveation of the stimuli, ensuring that the images are presented to one visual hemifield. The participants are required to indicate whether the image represented the right or the left hand by pressing the red or green button using the index and middle fingers of their unaffected hand. Speed and accuracy are both emphasised but there is no upper time limit for the response, and the button press initiates the next trial. Prior to the main task, participants complete a practice block of 12 trials (with 2000ms stimulus presentation times) that includes feedback, until they reach at least 75% accuracy across the entire practice block. They repeat the practice to ensure that they are able to perform the task above chance level. In the main task, there is a total of 100 trials (25 images × 2 hemifields × 2 depicted hands) conducted in a single block. Accuracy rates and RTs of the correct responses are calculated separately for the images of hands corresponding to the participant’s affected and unaffected limbs, and for the VFs corresponding to their affected and unaffected side of space (matched sides in healthy controls). As the task requires mental rotation of the images of hands, slower RTs and lower accuracy rates are considered to be an indicator of a distorted representation of the depicted limb [8, 80, 105]. To obtain single Hand Laterality Recognition indices, we will also calculate the differences in accuracy rates and RTs between depicted affected and unaffected hands. Positive accuracy index (unaffected – affected) and positive RT index (affected – unaffected) would indicate distorted representation of the affected limb.

**Figure 6.**
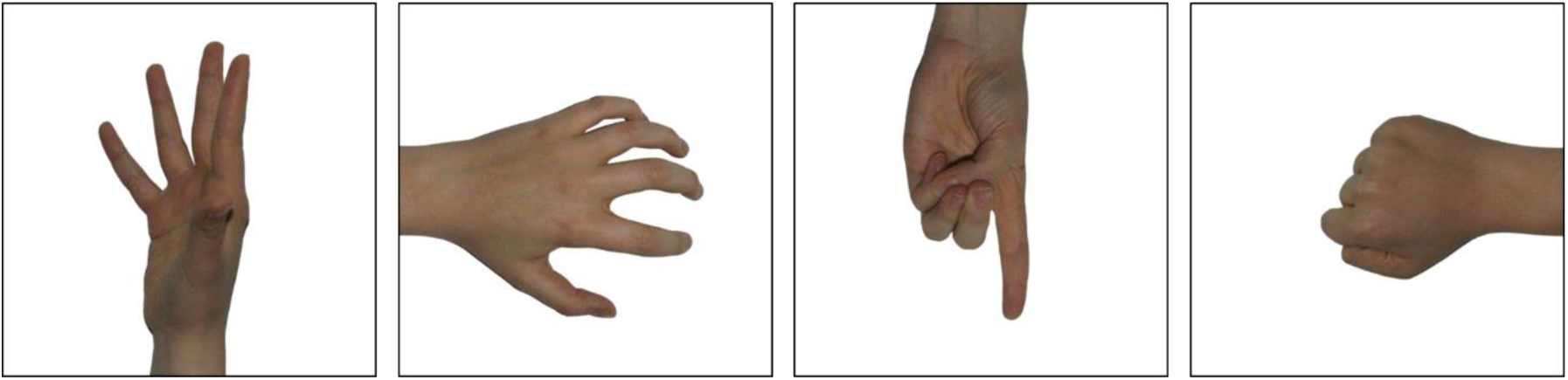
Example stimuli in the Hand Laterality Recognition task. Images of hands in four postures and rotation angles were included in the task.

### Blinding

All outcome measurements are recorded by a researcher who is blind to group allocations (MH). A researcher (JHB) who is not involved in data collection allocates participants with CRPS to treatment groups 1-5 days before RS2. JHB or another researcher not involved in data collection (ADV) trains the participants in how to carry out the PA treatment or sham treatment in a self-guided manner at the end of RS2. The participants return the goggles in sealed opaque bags after completing the treatment in RS3 so that the primary researcher (MH) remains blind to their treatment allocation. MH will be unblinded as to the group allocations of the participants once the last person has completed RS4, as there will be no further research sessions in which she will assess symptoms. Follow-up measurements in weeks 19 (LTFU1) and 31 (LTFU2) will be carried out via postal questionnaires scored by research assistants who are blind to the group allocations.

The participants will be blind to their group allocations as they are not made aware of the specific nature of the intervention beyond that it involves sensory-motor coordination, nor the type of goggles used in the other treatment arm. In the information sheet and the training materials, the same terms are used to describe both treatment arms. For instance, all participants will be informed that sensory-motor training involves reaching out to targets with their affected arm while wearing glasses fitted with lenses that distort vision. For ethical reasons, participants have to be told that they might receive either real or control treatment, and the meaning of double-blind randomised control trial will be explained to them in the information sheet and during training in how to carry out the treatment. A more general term “sensory-motor adaptation” is used to refer to PA in all study documents and instructions that the participants receive, to reduce the possibility that they could determine their treatment condition based on descriptions of PA on the Internet. At the end of the last in-person session (RS4) they will also be asked whether they have a belief about which condition they were allocated to, and their degree of confidence about this belief. They will be unblinded once data collection for this study is completed for all participants. Also, a participant might be unblinded before this time should they experience any worsening of symptoms that causes them concerns. If so, that participant will be withdrawn from the study in that no further data will be collected from them.

### Statistical analyses

To process and analyse the data we will use IBM SPSS Statistics [106], R [107], and MATLAB [108] software. Hypotheses will be tested using a significance level of *α* = .05. We will control type I errors in the primary analyses using Holm-Bonferroni corrections for multiple comparisons within each outcome analysis. No correction for multiple testing will be made in the exploratory analyses. We will report 95% bias-corrected and accelerated (BCa) bootstrap confidence intervals around all mean values.

Outliers are defined as scores outside ±3 SDs from the participant’s mean score for a task condition (participant-level data) or from the group mean (group-level data) for a particular test or task condition. We will examine participant-level and group-level RT data in the Hand Laterality Recognition and Directional Hypokinesia tasks for the presence of outliers and use nearest neighbour replacement if any are identified. We will use the same method of nearest neighbour replacement for the group-level outliers identified on the remaining outcome measures.

T-tests and ANOVAS will be conducted to compare mean values between groups and between data collection time points. Statistically significant interactions will be interrogated through follow-up contrasts. Wilcoxon signed-rank tests and Mann-Whitney U tests will be used if assumptions of t-tests are violated; however, ANOVAs are robust to moderate violations of normality and homogeneity of variance. Therefore, we will use ANOVAs unless severe violations of normality, homogeneity of variance, and sphericity assumptions are present, in which case we will use linear mixed models analyses with bootstrapping procedures.

#### Sample size and power calculation

The required sample size was calculated based on the primary outcome measure of self-reported pain [109]. A meta-analysis defined a clinically significant reduction in pain as a change of Δ = -2 (on a scale of 0-10) [110]. The sample size required to detect a pain reduction of this magnitude between RS2 and RS3 was estimated. The risk of type I error was set at 5% and the risk of type II error was set at 10%, giving 90% power to detect a significant change in the pain. The standard deviation expected in the current study was estimated as 1.98 based on pain intensity ratings obtained by our group in recent research [9]. Given these parameters, a minimum of 42 participants with CRPS (21 per group) is required to evaluate the effects of the PA treatment on the primary outcome measure of pain. Taking into account an anticipated drop-out rate of 20%, up to 52 participants with CRPS will be enrolled in order to obtain a total of at least 21 complete datasets for RS1-RS4 per treatment group. To provide normative data, 21 healthy (pain-free) control participants will be recruited.

#### Timing

No interim analyses are planned. The timing of the final analyses will be stratified by planned length of follow-up for the relevant outcome measures. Once all participants have completed RS4, we will analyse the RS1-RS4 data to address our research questions regarding the efficacy of PA treatment in reducing CRPS symptom severity (RQ1) and the relationships between the severity of clinical symptoms of CRPS and neuropsychological changes in perception of and attention to the affected limb and its corresponding side of space in RS1 (RQ5). We will conduct separate analyses of current self-reported pain intensity, the BPI, the Pain Detect Questionnaire, the BPDS, the Tampa Scale for Kinesiophobia, the Profile of Mood States, and the Patient’s Global Impression of Change scores once all participants meet the secondary endpoint (LTFU2). Recruitment will be terminated if we are not able to collect 42 full datasets for RS1-RS4 by 1 March 2019.

#### Treatment outcome analyses

We will conduct intention to treat (ITT) as our primary analysis to examine the overall effects of prism adaptation. The ITT analysis will include all allocated participants with CRPS regardless of treatment adherence and completion of outcome measurements. Baseline post-randomisation observation (RS2) carried forward will be used to account for missing data in ITT analysis, as the participants are expected to return to pre-treatment baseline over time. The exception is the Patient Global Impression of Change questionnaire that is only completed in the post-treatment research sessions, in which case the RS3 observation will be carried forward. Missing data from the computer-based tasks within each research session will be replaced by group mean for the particular task condition (with an exception of Directional Hypokinesia task, where some participant with CRPS may not be able to complete the task using the affected limb; in such cases only the conditions completed with the unaffected limb will be analysed). Missing daily logbook ratings will be interpolated using linear regression.

We will also conduct a supportive per-protocol (PP) analysis to see whether PA treatment can benefit the participants with CRPS who were able to perform it according to the trained protocol compared to those participants who were able to complete the sham treatment according to trained protocol [111]. The PP population will be the subset of the ITT population who provided complete outcome data for RS1-RS4 (i.e. attended all in-person research sessions and completed the primary outcome measures) and missed no more than 6 treatment sessions.

We will use confidence intervals to compare the RS1 primary outcomes scores of the participants with CRPS who withdrew and those who remained in the trial until RS4 to assess any potential selection bias. The timing and reasons for withdrawal will be presented in a CONSORT diagram.

#### Descriptive characteristics

We will report baseline characteristics for individual participants with CRPS such as affected limb, type of inciting injury, CRPS duration, co-morbidities, prescribed medications and other treatments, and change in handedness score.

Minimisation factors listed in Table 2 will be presented as group means for continuous factors or proportion of participants in each group who are classed positive on each categorical factor. We will conduct a series of contrasts and chi-square tests to confirm that the minimisation procedure successfully equated the two groups on these factors. Contrasts will also be used to confirm that the PA and sham treatment groups are matched on mean Profile of Mood States, Tampa Scale for Kinesiophobia, Revised Life Orientation Test, and Patient Centred Outcomes Questionnaire scores.

#### Efficacy of PA treatment in reducing pain and CRPS symptom severity (RQ1)

A 2×6 ANOVA with Group as a between-subjects factor (PA treatment, sham treatment), and Time (RS1, RS2, RS3, RS4, LTFU1, LTFU2) as a within-subject factor will be conducted for the first primary outcome of pain intensity rating. We will also conduct sixteen *a-priori* contrasts to compare RS1 vs. RS2, RS2 vs. RS3, RS3 vs. RS4, RS2 vs. RS4, RS2 vs. LTFU1, RS4 vs. LTFU1, LTFU1 vs. LTFU2, and RS2 vs. LTFU2 within each group. We will also conduct a 2×4 ANOVA with the factors Group (PA treatment, sham treatment) and Time (RS1, RS2, RS3, RS4) for the second primary outcome of CPRS severity score, followed by eight *a-priori* contrasts comparing RS1 vs. RS2, RS2 vs. RS3, RS3 vs. RS4, and RS2 vs. RS4 within each group. We are primarily interested in detecting any changes between RS2 and RS3 which would represent immediate effects of treatment.

Minimisation factors (see Table 2) may be included as covariates in the ANOVAs if there are significant differences at RS1. Similarly, if we find significant group differences in the Profile of Mood States, Tampa Scale for Kinesiophobia, or Revised Life Orientation Test RS1 scores, these variables may be used as covariates in the ANOVAs on pain and CRPS severity score.

We will also calculate the Number Needed to Treat (NNT) separately for pain and CRPS severity score. The NNT will be based on the proportion of participants in each treatment arm that achieved clinically significant pain relief (≥2 points on 0-10 NRS scale [110]) and reduction in CRPS symptom severity (≥4.9 points [43]) in RS3 compared to RS2.

#### Effects of PA treatment on neuropsychological changes and other secondary outcomes (RQ2) and time course of any improvements (RQ3)

To analyze between-group (PA treatment vs. sham treatment) differences on the secondary outcome measures (see Table 3) across four (RS1-RS4) or six (RS1-LTFU2) time points, we will conduct 2×4 or 2×6 ANOVAs as described for the analyses of the primary outcomes. Specifically, a 2×4 ANOVA will be run on each clinical assessment outcome (limb temperature asymmetry, hands size difference, grip strength and ΔFTP ratios, MDT, MPT, two-point discrimination threshold ratios, and allodynia) and mean group scores in the following computer-based measures: PSSs in the TOJ task, PSEs in the Landmark task, attention bias scores in the Greyscales task, and bias scores in the MNLB task. We will also use 2×4 ANOVAS to analyse between-group differences on indices A and B for the affected and unaffected hand performance in the Directional Hypokinesia task, as well as on hand laterality recognition accuracy and RTs indices in the Hand Laterality Recognition task across RS1-RS4. Separate 2×6 ANOVAs will be run on mean group scores on each of the self-reported questionnaire measures: pain intensity and interference components of the BPI, the Pain Detect Questionnaire, The BPDS, the Tampa Scale for Kinesiophobia, and the Profile of Mood States.

We will plot group means of daily ratings of average pain, range of movement, and interference over time and use contrasts to identify the time points of significant group differences. We will also identify for both groups and for each measure the average number of days to reach peak improvement from the start of treatment, and the average number of days from the peak improvement to return to baseline.

#### Predictors of the response to PA treatment and / or CRPS progression over time (RQ4)

We will conduct exploratory analyses of the potential factors that can predict response to treatment for the PA group. First, we will calculate reduction scores as a change in current pain scores and CRPS severity scores from the immediate pre-treatment to immediate post-treatment sessions (RS3 – RS2). Second, we will conduct two separate linear mixed models regressions on pain reduction scores and CRPS severity reduction scores including the pertinent explanatory factors such as demographic characteristics; current pain intensity; CRPS severity score; and scores on the self-report questionnaires, clinical assessments, and computer-based tests. In the first instance, we will consider those outcomes that differed the most from the healthy control participants in RS1 (see statistical analyses for RQ5 in the next section).

The same factors will be considered potential explanatory variables in linear mixed models regressions on current pain scores and CRPS severity scores across four research sessions (RS1-RS4). These exploratory analyses will be conducted for data from all the participants with CRPS to examine possible predictors of CRPS progression over time (including but not limited to treatment group).

#### Baseline neuropsychological symptoms and their relationships with the clinical symptoms of CRPS (RQ5)

We will conduct a series of contrasts to compare mean age, proportion of males and females, and proportion of left- and right-handed individuals between participants with CRPS in RS1 (total CRPS sample, regardless of subsequent treatment allocation) and healthy controls. Participants’ mean scores on self-report questionnaires and clinical assessments in RS1 will also be compared between the two groups. Specifically, we will conduct contrasts to compare participants with CRPS and healthy controls groups on the BPDS and Profile Of Mood States scores; the hand laterality indices (current for healthy controls, and handedness before CRPS for the participants with CRPS); limb temperature asymmetry and hands size difference (affected – unaffected), grip strength and ΔFTP ratios (affected / unaffected), MDT, MPT, two-point discrimination threshold ratios, and mean allodynia score on the affected side.

To test whether the participants with CRPS in RS1 show visuospatial attention bias compared to healthy controls, we will use four separate contrasts. Specifically, we will conduct four between-group comparisons of the following variables: PSSs in the TOJ task, PSEs in the Landmark task, attention bias scores in the Greyscales task, and bias scores in the MNLB task.

The Directional Hypokinesia task conditions performed with the affected and unaffected hand will be analyzed separately. After excluding incorrect and missed trials, we will use mean movement initiation times (MITs) and movement execution times (METs) for each combination of VF in which the target appeared and hand Starting Position to test if the participants with CRPS show signs of directional hypokinesia compared to the healthy controls. We will conduct two three-way ANOVAs on MITs for each hand with the following factors: Group (participants with CRPS, healthy controls), VF (affected, unaffected), and Starting Position (affected, central, unaffected). Significant interactions will be followed by four *a-priori* contrasts to test whether the participants with CRPS are slower to initiate movements toward the targets in their affected side of space (regardless of the direction of reaching movement required) and / or in the direction toward their affected side of space (regardless of the location of the target). Specifically, we will examine if participants with CRPS have slower MITs to the targets in the affected VF than in the unaffected VF; to the affected VF compared to healthy controls; to the affected VF from central compared to affected Starting Position; and to the affected VF from central Starting Position compared to healthy controls. Analogous analyses will be conducted on METs to test if participants with CRPS show signs of directional bradykinesia compared to healthy controls. We will also examine differences between Groups (participants with CRPS, healthy controls) on MITs and METs through separate contrasts for each index (A and B) of directional hypokinesia and bradykinesia. As further exploratory analyses we will examine how many participants with CRPS are impaired on both indices (A and B) of directional hypokinesia and bradykinesia by identifying which participants obtained positive A and B indices and by comparing each CPRS patient’s indices to the mean indices for healthy controls using Crawford t-tests [112].

To test for differences in body representation as measured by the Hand Laterality Recognition task between the participants with CRPS and healthy controls, we will conduct two three-way ANOVAS with the factors Group (participants with CRPS, healthy controls), depicted Hand (affected, unaffected), and VF (affected, unaffected) on accuracy rates and RTs to accurate responses. If there are significant three-way interactions, we will conduct *a-priori* contrasts to test whether the participants with CRPS are less accurate and / or slower in responding to: the depicted hands corresponding to their affected hand compared to those corresponding to their unaffected hand when the hands are presented in the affected VF; the affected hands presented in the affected VF compared to the unaffected VF; the affected hands compared to the unaffected hands; the affected hands presented in the affected VF compared to healthy controls; the affected hands compared to healthy controls; and the hands presented in the affected VF compared to healthy controls. If there is no effect of VF, we will only consider accuracy rates / RTs to recognize affected and unaffected hands averaged across both VFs in follow-up contrasts and any further correlation / regression analyses.

### Relationships between neuropsychological changes and clinical symptoms of CRPS

Correlation and regression analyses will be conducted to test relationships between neuropsychological changes (as measured by computer-based tasks) and clinical symptoms of CRPS (as measured by self-report questionnaires and clinical assessments). These analyses will depend on which outcomes show significant differences between participants with CRPS and healthy controls, thus they are exploratory.

## Discussion

Considering the poor overall response to conventional medical treatments for CRPS [113], it is important to seek novel methods of pain relief and symptoms improvement. PA is an emerging treatment that targets spatial attention deficits, has shown early promise as an intervention for CRPS, and might operate through different mechanisms to mirror visual feedback (another neurocognitive treatment for CRPS [23, 114]).

The ongoing trial that is described in this protocol is the first to investigate the effects of PA treatment on pain and CRPS symptom severity using a double-blind, randomized, and sham-controlled design in a patient sample that is large enough to detect a clinically significant reduction in pain. These aspects of our design, as well as stratified randomisation and intention to treat and per-protocol analyses will allow unbiased evaluation of a brief, low-cost treatment that can be easily self-administered by the participant in a home setting.

However, self-guided administration of the treatment might also be its limitation in the resent study, as it prevents us from directly monitoring participants’ compliance. This is considered a necessary trade-off to test the treatment as it would be most likely integrated into CRPS management. Furthermore, opting for home-based treatment will aid the recruitment of a sufficient number of participants, who are drawn from a broad geographical area, as it would not be feasible for them to travel to the research centre for each treatment session, or for another researcher to repeatedly assess their compliance. We put in place several measures to encourage adherence to treatment, such as in-person training, instructions and guidance in multiple media, and easy access to advice. To avoid adding another layer of difficulty and increasing potential burden of treatment (especially for those participants who do not have very good technical competence), we decided against asking participants to video-record their treatment sessions. Thus, we rely solely on self-reported adherence, that is, recording the timing and completion of each treatment session in daily logbooks. This could limit the interpretation of our findings; however, we will evaluate them considering the possibility that participants may have not complied with the treatment regimen as expected.

While clearly defining our primary outcome measures, the trial includes a wide range of secondary outcome measures to assess the impact of treatment on pain and other clinical CRPS symptoms, as well as neuropsychological and emotional functioning, and symptoms’ interference with daily life. These measures will allow us to explore relationships between self-reported, clinical, and neuropsychological manifestations of CRPS from baseline data, independent of the primary research aim of testing the efficacy of PA.

Despite their informative value, large number of secondary outcomes might reduce the quality of data. To mitigate the potential impact of long duration and burden of research sessions, we composed a battery of assessments that would take no longer than four hours to complete, including breaks between assessments. We also provide the participants with overnight accommodation near the research centre in cases when long travels are required, to minimise their fatigue during research sessions. In light of how little is known about cognitive changes in CRPS and effects of PA on these changes, broad battery of neuropsychological tests seems appropriate.

Furthermore, this research may identify potential individual differences accounting for the course of CRPS and response to treatment. The findings could provide an indication of how to identify the patients who are most likely to benefit from PA based on their cognitive and physical symptoms. This would inform subsequent research and therapies.

If PA brings benefits beyond that of the sham treatment on the primary outcome measures of the ongoing trial, this treatment should be developed as a recommended method to reduce pain and other CRPS symptoms. The study is likely to expand on our limited understanding of this debilitating condition and its neuropsychological components.

## Data Availability

There is no data associated with the current paper, which describes a protocol for a clinical trial that is in progress at the time of submission. All investigators will have access to the final dataset. Anonymised participant-level data from the trial will be stored using the Open Science Framework repository and access will be granted upon request. Access shall be requested by contacting the corresponding author via email. Description of the data and specific instructions for gaining access will be listed in a publicly-available format in the repository. The data will be made available in this format at the time that the paper describing the trial outcome is published, upon MH’s graduation from her PhD, or two years after the end of data collection (whichever comes first).

## List of abbreviations

BPDS: Bath CRPS Body Perception Disturbance Scale
BPI: Brief Pain Inventory
CRPS: Complex Regional Pain Syndrome
FTP: Finger-To-Palm distance
LTFU1: Long Term Follow-Up 1 (week 19) by post
LTFU2: Long Term Follow-Up 2 (week 31) by post
MDT: Mechanical Detection Threshold
MET: Movement Execution Time
MIT: Movement Initiation Time
MPT: Mechanical Pain Threshold
NRS: Numerical Rating Scale
PA: Prism Adaptation
PSE: Point of Subjective Equality
PSS: Point of Subjective Simultaneity
RS1: Research Session 1 (week 1)
RS2: Research Session 2 (week 4)
RS3: Research Session 3 (week 7)
RS4: Research Session 4 (week 11)
RSDSA: Reflex Sympathetic Dystrophy Syndrome Association
RT: Reaction Time
QST: Quantitative Sensory Testing
TOJ: Temporal Order Judgement
VF: Visual Field

## Declarations

### Ethics approval and consent to participate

This trial gained approval from National Health Service Oxfordshire Research Ethics Committee A and Health Research Authority, reference number 12/sc/0557, and from the University of Bath Psychology Ethics Committee (16-333). Major protocol amendments are implemented only after formal approval from the Research Ethics Committee. Any protocol updates are also documented and dated on the ISRCTN trial record. The researcher who is responsible for data collection (MH) obtains informed consent from each participant that is signed and dated prior to any study-related procedures. Verbal consent to continue with the study is taken at the beginning of each consecutive research session. Participants are able to withdraw from the trial at any time.

The study sponsor, the University of Bath, is involved in the trial management through providing data storage and ethical supervision (Contact: Prof. Jonathan Knight, Claverton Down Road, Bath BA2 7AY, UK).

### Consent for publication

Not applicable.

### Availability of data and material

There is no data associated with the current paper, which describes a protocol for a clinical trial that is in progress at the time of submission. All investigators will have access to the final dataset. Anonymised participant-level data from the trial will be stored using the Open Science Framework repository and access will be granted upon request. Access shall be requested by contacting the corresponding author via email. Description of the data and specific instructions for gaining access will be listed in a publicly-available format in the repository. The data will be made available in this format at the time that the paper describing the trial outcome is published, upon MH’s graduation from her PhD, or two years after the end of data collection (whichever comes first). Obtained results will be communicated to participants, clinicians, and the public through newsletters, talks and press releases.

Materials that were modified or developed specifically for the purpose of the current trial, as well as data management plan, will be available from the project’s webpage at Open Science Framework upon completion of data collection for the trial.

## Authors’ contributions

The study was conceived of by JHB. JHB, MH, ADV and MJP contributed to design. JHB and MJP secured the funding. MH developed the testing protocols, programmed the computer-based tasks, and prepared the draft. MH and JHB devised the randomisation with minimisation procedure. All the authors have read and approved this version of the manuscript.

### Acknowledgements

Not applicable.

## Additional files

Additional file 1.docx – Supplementary Table 1 lists all items from the World Health Organisation Trial Registration Data Set.

Additional file 2.docx – Supplementary text describing the procedure and results of a pilot study to select the stimulus set for the Hand Laterality Recognition task.

## Footnotes

1 Note that we will not exclude any participants based on having large fluctuations in symptoms between two baseline assessments (RS1 ad RS2), if they meet the CRPS diagnostic research criteria (see “Eligibility criteria” section).

2 We will not exclude participants who have CRPS in ipsilateral lower limb if the upper limb is the primarily affected site and pain and other symptoms are not less severe than in the lower limb. Those participants, as well as participants with diagnoses of other chronic pain conditions (as long as these are less severe than CRPS), will complete the relevant self-reported outcome measures (i.e. questionnaires about pain) separately for the primary CRPS-affected upper limb, and separately for other chronic pain. We will measure the primary outcome of CRPS symptom severity only for the upper limb. Anecdotally, CRPS participants previously studied by our research group can easily differentiate CRPS pain and other symptoms in one extremity from another, and from other chronic pain conditions. Our primary analyses will only concern the pain and CRPS severity data regarding the CRPS-affected upper limb, however, data related to other pain might be used in exploratory analyses.

